# The Relationship Between Moderate-to-Vigorous Physical Activity and Hearing Health in the UK Biobank

**DOI:** 10.64898/2026.01.14.26343944

**Authors:** Bridget N. Carey, Timothy P. Morris, Joseph M. Northey, Charles H. Hillman, Jonathan E. Peelle

## Abstract

**Objective:** Because hearing difficulties contribute significantly to years lived with disability and global economic burdens, finding ways to support hearing health is crucial. A key challenge is understanding how to improve speech perception in the presence of background noise, which is a significant complaint among listeners with and without hearing loss. Understanding speech in noise requires more than simply hearing the conversation but rather also demands cognitive processes such as memory for what was said and selective attention to the target speech instead of competing noise. Because physical activity has been shown to benefit cognitive health more broadly, we surmised it may have a positive relationship with hearing health. We aimed to investigate the association between time spent in moderate-to-vigorous physical activity and self-reported hearing problems in a comprehensive population dataset.

**Design:** This was a prospective cohort study where participants (N= 79,286; 54.57% female) in the UK Biobank dataset wore an accelerometer for 7 days and completed questionnaires about self-reported hearing problems. After dividing participants into two groups based on their answers to hearing health questions, we ran a logistic regression to determine if time spent in moderate-to-vigorous physical activity decreases the likelihood of reporting hearing difficulties generally, or specifically in the presence of background noise. In addition, we investigated whether spending at least 150 minutes in moderate-to-vigorous physical activity, corresponding to the World Health Organization physical activity guidelines, was associated with hearing outcomes.

**Results:** Spending more time in moderate-to-vigorous physical activity was associated with lower odds of reporting a problem with hearing (OR=0.990, 95% CI [0.983, 0.997], p=0.005) and lower odds of reporting challenges understanding speech in noise (OR= 0.991, 95% CI [0.985, 0.998], p= 0.007). Additionally, meeting physical activity guidelines was associated with lower odds of reporting both problems with hearing (OR= 0.958, 95% CI [0.923, 0.995], p= 0.025) and problems understanding speech in noise (OR= 0.953, 95% CI [0.922, 0.986], p= 0.005).

**Conclusions:** These findings suggest that spending more time in moderate-to-vigorous physical activity is associated with better hearing health, specifically lower odds of reporting problems with hearing or understanding speech in noise. Targeting physical activity as a non-invasive and low-cost behavioral intervention may make the common issue of hearing loss more manageable.

## Introduction

Hearing loss has often been characterized as an “invisible disability” leading to miscommunications, exclusion, and barriers to treatment (Aldè et al., 2025). As the world’s population ages, by 2050 close to 2.5 billion people will experience some form of hearing loss (World Health Organization, 2021). In addition to adversely affecting social interactions (Shukla et al., 2020), cognitive performance (Lin et al., 2011; Loughrey et al., 2018; Powell et al., 2022; Taljaard et al., 2016) and mental health (Golub et al., 2020; McClannahan et al., 2025), unaddressed hearing issues cost around $1 trillion globally each year (Ghebreyesus, 2024). Thus, there is a critical need to support hearing health throughout the lifespan and investigate how it affects healthy aging. In the current study we focus on two complementary aspects of hearing health. The first is overall hearing difficulties, primarily reflecting the integrity of the peripheral auditory system highlighting cochlear integrity. The second is the ability to understand speech amid background noise, which also relies on the central auditory system and requires higher-level cognitive resources beyond just detecting sound.

Outside the realm of hearing health, there is a long history of research highlighting the benefits of physical activity on multiple facets of healthy aging (Blair & Morris, 2009). Being active reduces the risk of several chronic diseases such as cardiovascular disease and obesity and increases longevity (Miller et al., 2016). More physical activity also results in better overall well-being and mental health (Kim et al., 2012), moderates stress systems (Anderson et al., 2013; De Nys et al., 2022; Duclos & Tabarin, 2016; Fragala et al., 2011), is correlated with improved sleep quality (De Nys et al., 2022; Kredlow et al., 2015; Vanderlinden et al., 2020; Wang & Boros, 2021), and is positively associated with overall general health factors (Dhuli et al., 2022). Although the mechanisms underlying these beneficial relationships are not fully understood, they likely encompass changes at molecular, cellular, and systems levels such as protecting musculoskeletal function during aging (Neufer et al., 2015). These same constellation of effects may extend to hearing health.

There is some evidence regarding the association between physical activity, specifically moderate-to-vigorous physical activity (MVPA), and hearing difficulties. Older adults with hearing loss are less physically active (Assi et al., 2024) and decrease physical activity levels faster than those without hearing loss (Goodwin et al., 2023). Specifically, older adults with hearing loss spend less time in MVPA than those with normal hearing (Kuo et al., 2021). Other researchers have investigated this relationship in the opposite direction such that more time in exercise leads to reduced risk of hearing loss and improved pure-tone thresholds (Li et al., 2025). Some have found that being more active or spending more time in MVPA is associated with better pure-tone audiometry thresholds (Haas et al., 2016; Kolkhorst et al., 1998; Loprinzi et al., 2014) and lower odds of having a hearing impairment (Gispen et al., 2014).

In a more causal manner, longitudinal studies have shown that being more physically active may be a potential protective factor for mitigating the risk of developing future hearing loss where higher levels of physical activity are shown to lower incidences of developing hearing loss as one ages (Curhan et al., 2013; Kawakami et al., 2022). Those who meet physical activity guidelines, and therefore spend more time in MVPA, report better hearing thresholds than those who do not (Martinez-Amezcua et al., 2023). While further longitudinal work is needed to confirm the directionality of this relationship, the current study expands upon these findings.

A second facet of hearing health addressed in this study is the ability to understand speech in noisy environments. To understand speech in noise, a listener must be able to hear the words spoken, hold in memory what was said, and process the meaning behind words in the context of sentences (Pichora Fuller et al., 1995). Collectively, these actions tap into cognitive processes such as working memory and selective attention, often called executive functions (Miyake et al., 2000). The additional challenge associated with understanding speech in noise suggests that understanding speech in noise increases cognitive load which forces listeners to rely on resources outside the peripheral auditory network (Mattys et al., 2025; Peelle, 2018; Wingfield et al., 2005). Consistent with this hypothesis, understanding acoustically challenging speech consistently requires an upregulation of executive functions and increased activity in the frontal cortex (Davis & Johnsrude, 2003a; Gosselin & Gagné, 2011; Sherafati et al., 2022; Wild et al., 2012; Wong et al., 2009). If understanding speech in noise relies on executive functions, practices that have been validated to boost executive function and the associated brain networks that support them might improve listeners’ understanding of speech in noise.

To our knowledge there are no studies directly examining the relationship between MVPA and speech-in-noise understanding. However, there is a rich history of research elucidating the benefits of physical activity on executive function. Compared to sedentary older adults, meeting MVPA guidelines presented by the World Health Organization of at least 150 minutes per week (World Health Organization, 2026) is associated with improved performance on executive function tasks (O’Brien et al., 2021). There is also moderate evidence of an association between MVPA and improvements in cognition, neuropsychological assessments, executive function, processing speed, and memory (Erickson et al., 2019; Van Praag, 2009). Additionally, greater amounts of physical activity are associated with increased neural efficiency during cognitively challenging tasks (Erickson et al., 2015) and following exercise there is increased activity in frontal and parietal regions (Won et al., 2019). Beyond proximal effects there may also be protective effects present in older age related to a lifetime of physical activity habits such as prevention or delayed loss of cognitive decline and neurodegenerative diseases (Dik et al., 2003; Van Praag, 2009).

In summary, while the most common complaint from those who are treated for hearing loss comes from difficulty understanding speech in background noise (Billings et al., 2024; Kochkin, 2010), no comprehensive study has directly explored its association with MVPA. Given that speech-in-noise understanding relies on these same processes that are positively correlated with physical activity, we aimed to examine whether time spent in MVPA would be associated with speech-in-noise problems. The current study also aims to add to the literature investigating the relationship between hearing problems and MVPA using objective accelerometer data in a population-based cohort. Understanding these gaps is critical to managing the myriad burdens that come with hearing impairments in the general population. Thus, we evaluated whether spending more time in MVPA was associated with lower odds of reporting hearing difficulties and speech-in-noise problems in a sample of 79,286 participants from the UK Biobank.

## Materials and Methods

Access to UK Biobank data is restricted to those with an approved project. Analysis code for the current project is available at https://osf.io/djn5m/files.

### Design and Participants

The UK Biobank is on ongoing prospective cohort study of approximately half a million volunteer participants across the United Kingdom. Baseline recruitment occurred from 2006-2010 when participants (n=503,317) were between 40 and 69 years old (Sudlow et al., 2015). Participants were tested at 22 different assessment centers and covered diverse socioeconomic and ethnic backgrounds. Initial recruitment occurred through the mail from the National Health Service (NHS) central registers. Detailed information was collected at baseline assessment including lifestyle measures, physical measures, and biological specimens. Selection criteria for the current study are shown in Figure 1. Of the total participants, 502,128 had complete baseline data and 469,592 had complete hearing data. Between February 2013 and December 2015, a subset of participants (n=103,648) was randomly invited and subsequently wore an accelerometer for 7 days to objectively measure physical activity. Of the people with valid hearing data, 79,286 also had valid accelerometer and covariate data and comprised our final sample. Data were obtained through a UK Biobank application (ID 140089). Participants provided signed informed consent and the research was approved by the North West Multi-Centre Research Ethics Committee (Sudlow et al., 2015).

**Figure 1.**
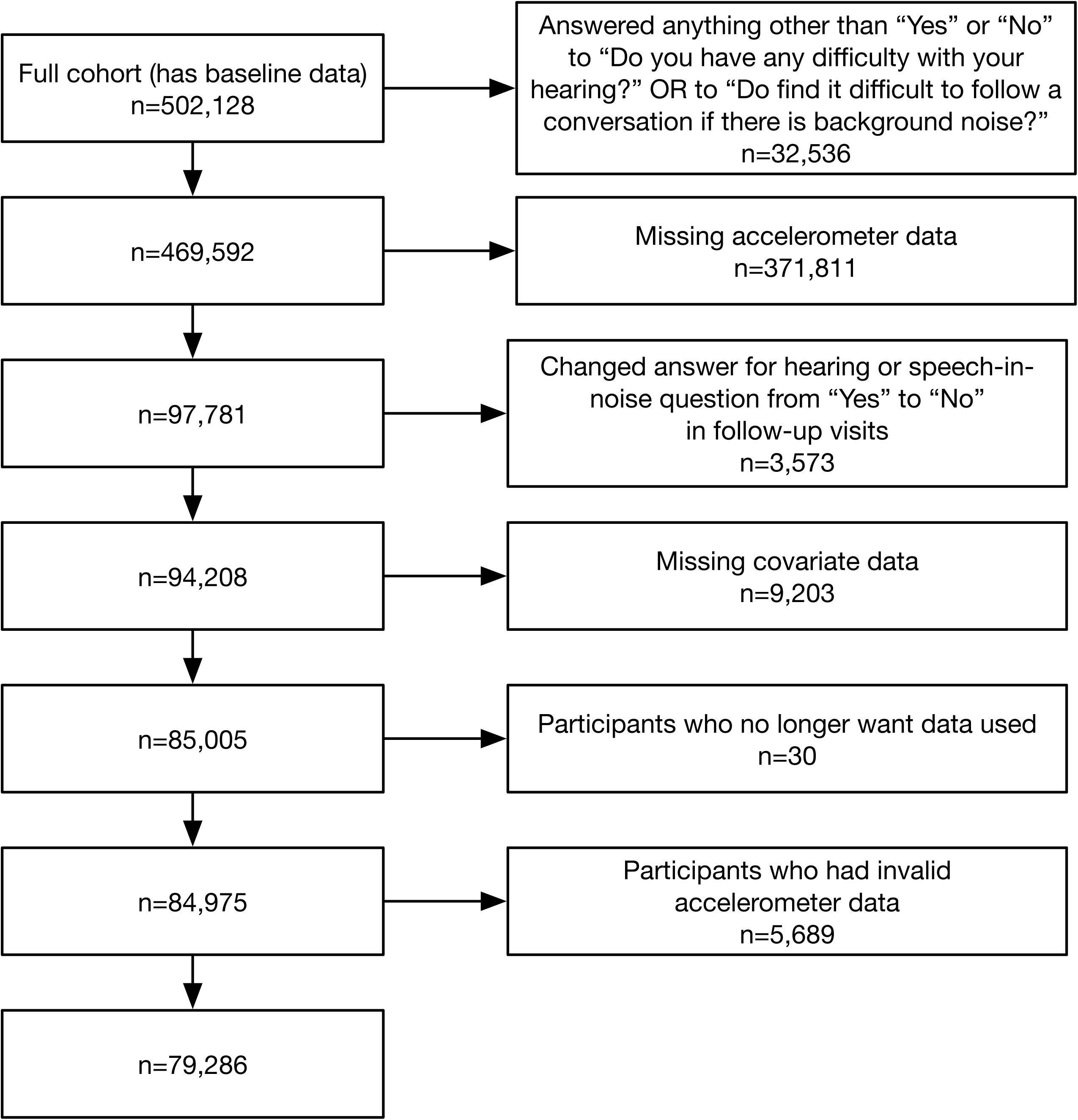
Diagram illustrating the process of selecting participants from the UK Biobank dataset.

### Hearing and Speech-in-Noise Outcomes

Hearing outcome came from the touchscreen questionnaire question, “Do you have any difficulty with your hearing?” (Data-Field 2247). The speech-in-noise outcome came from the touchscreen questionnaire question, “Do you find it difficult to follow a conversation if there is background noise (such as TV, radio, children playing?” (Data-Field 2257). Only “Yes” or “No” answers to these questions were included in the analyses. Other options included “I am completely deaf”, “Do not know”, and “Prefer not to answer.” Participants were also excluded if they answered “Yes” to a question at baseline assessment but answered “No” in follow-up sessions.

### Physical Activity

Between February 2013 and December 2015, participants who had provided a valid email address were randomly selected and sent an email invitation to wear an accelerometer (Axivity AX3 wrist-worn triaxial accelerometer) for 7 days (Doherty et al., 2017). Participants were told to wear the accelerometer continuously on their dominant wrist while conducting their normal activities. Processed physical activity data is provided by the UK Biobank (Walmsley et al., 2022), which uses a machine learning model to estimate time in MVPA (Data-Field 40049). As per previous work with this dataset, we excluded participants if they did not have at least 3 days of data and data in each 1-hour period of the 24-hour cycle (Data-Field 90015).

### Covariates

We chose covariates based on current knowledge of possible confounders and included age at baseline (Data-Field 21022), sex (Data-Field 31), education level (Data-Field 6138), income (Data-Field 738), ethnicity (Data-Field 21000), alcohol use (Data-Field 20117), smoking (Data-Field 20116), diabetes (Data-Field 2443), vascular/heart problems (Data-Field 6150), and body mass index (BMI) (Data-Field 21001). Education was divided into higher and lower education based on if the participant had at least a college/university degree or another professional qualification. Income was dichotomized as high or low (< £51,999 and > £52,000). As the vast majority of the UK Biobank sample is White, ethnicity was dichotomized as White and Non-White. Alcohol use and smoking were categorized as never, current, previous use, or prefer not to answer. Diabetes answers included, yes, no, don’t know, and prefer not to answer. Vascular/heart problems were dichotomized as yes or no (if they had a heart attack, angina, stroke, or high blood pressure). We also created a time difference covariate to account for the time between the baseline visit and accelerometer wear period.

### Statistical Analyses

To characterize baseline measurements, we summarize continuous variables as means with standard deviations (SDs), and categorical variables by count (percentages). We used multivariable logistic regressions to examine the association between MVPA and hearing and speech-in-noise outcomes. To make the results more interpretable, we also scaled accelerometer-based MVPA data by 100 so that a 1-unit increase was equivalent to a 1% increase in time spent in MVPA. All models controlled for age at baseline, sex, education level, income, ethnicity, alcohol use, smoking, diabetes, vascular/heart problems, and BMI. Age was mean-centered. Model 1 examined the odds of reporting a hearing problem given the percentage of time participants spent in MVPA accounting for the covariates. Model 2 was similar except the outcome variable was reported speech-in-noise problems instead of hearing problems.

In Models 3 and 4, we wanted to investigate whether meeting World Health Organization guidelines of 150 minutes per week in MVPA affected the odds of reporting a hearing or speech-in-noise problem, respectively, while accounting for covariates. All models passed assumption checks except for linearity of logit odds assumption. Models with hearing as the outcome variable (Models 1 and 3) had BMI as a predictor violating this assumption and models with a speech-in-noise problem as an outcome variable (Models 2 and 4) had BMI and age violating this assumption. We modeled these predictors using restricted cubic splines (df = 4) to include nonlinear relationships, but the results did not change. For simplicity, we report the results for the models without restricted cubic splines but include the spline models in Supplemental Material (Supplemental Tables 1-4). All statistical analyses were performed using RStudio 4.4.0 (R Core Team, 2024) on UK Biobank’s DNAnexus platform. Values of p <.05 were considered significant. Data analysis was performed from November 2023 to September 2025.

## Results

### Participant Baseline Characteristics

The mean (SD) age of participants was 55.95 (7.86). Out of the total 79,286 participants, 54.57% of the participants were female, 96.88% were White, 24.26% reported having hearing problems, and 34.50% reported having speech-in-noise problems. Compared to those that said they did not have hearing problems (M = 2.94%, SD = 2.43%), those who reported hearing problems spent less time in MVPA (M = 2.87%, SD = 2.45%) by 7.05 minutes per week (296.35 minutes per week versus 289.30 minutes). The difference was found to be statistically significant, *t*(32,194) = 2.41, *p* < .01 but the effect size was negligible (*d* = 0.028). There was also a significant difference in time spent in MVPA for those who reported a speech-in-noise problem (M=2.90%, SD=2.45%) versus those who did not (M=2.93%, SD=2.43%), *t*(55,254) = 1.76, *p* = .040 (292.32 minutes versus 295.34 minutes) but the effect size was negligible (*d* = 0.013).

### Associations of Hearing and Speech-in-Noise Outcomes with Time Spent in MVPA

After adjusting for covariates in the Model 1, spending more time in MVPA was significantly associated with lower odds of reporting a hearing problem (OR=0.990, 95% CI [0.983, 0.997], p=0.005) as shown in Table 1. This means that for every 14.4 minutes more a person spends in MVPA per day (or 100.80 minutes per week), the odds of reporting a hearing problem decrease by 1.00%. Increasing time spent in MVPA to 72 minutes a day (or 504 minutes per week) decreases the odds of reporting a hearing problem by 4.90%. In Model 2, after adjusting for these covariates, spending more time in MVPA was also significantly associated with lower odds of reporting a speech-in-noise problem (OR= 0.991, 95% CI [0.985, 0.998], p= 0.007) as shown in Table 1. This means that for every 14.4 minutes more a person spends in MVPA per day (or 100.80 minutes per week), the odds of reporting a hearing problem decrease by 0.90%. Increasing time spent in MVPA to 72 minutes a day (or 504 minutes per week) decreases the odds of reporting a hearing problem by 4.42%.

**Table 1.**
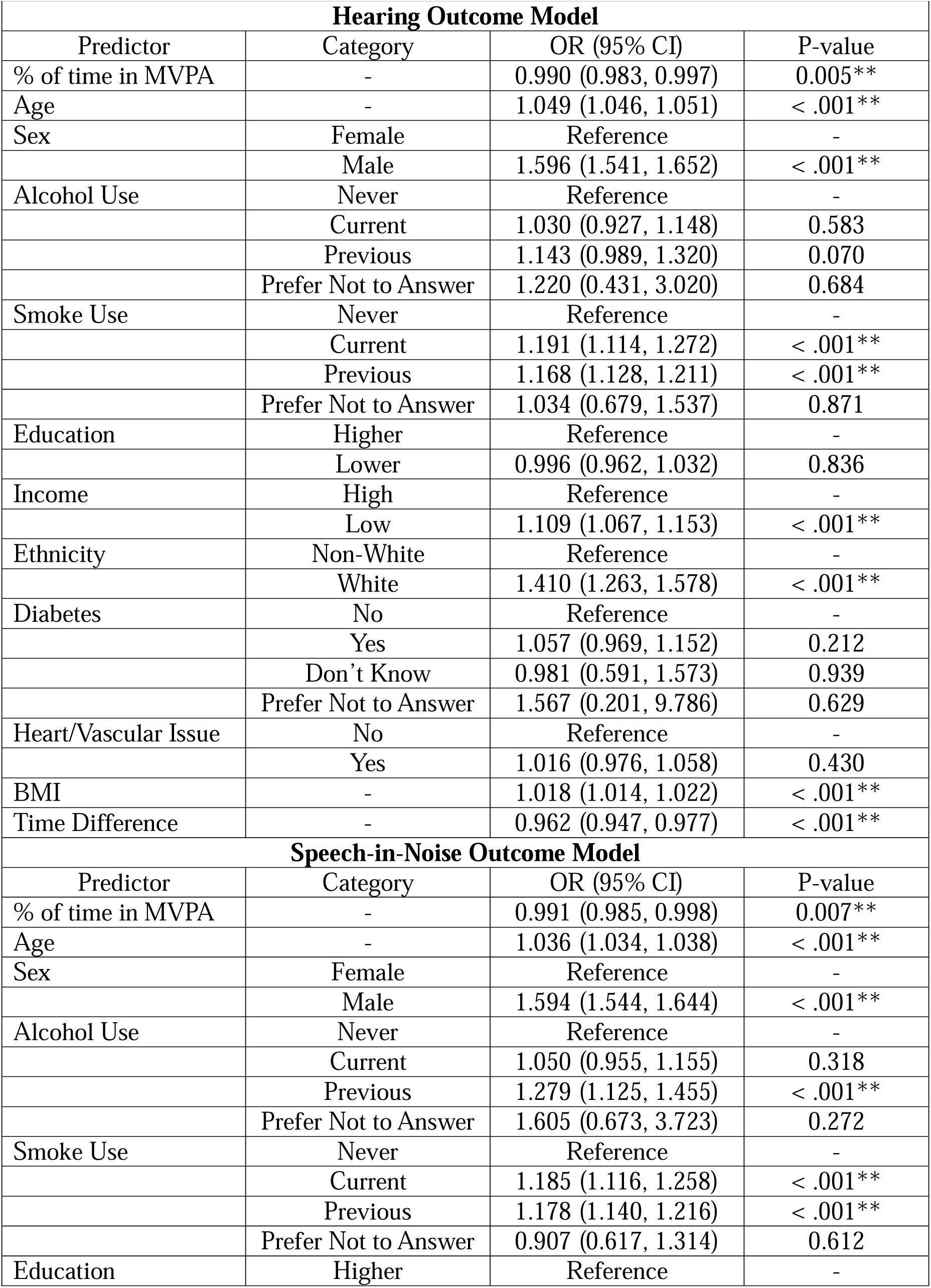
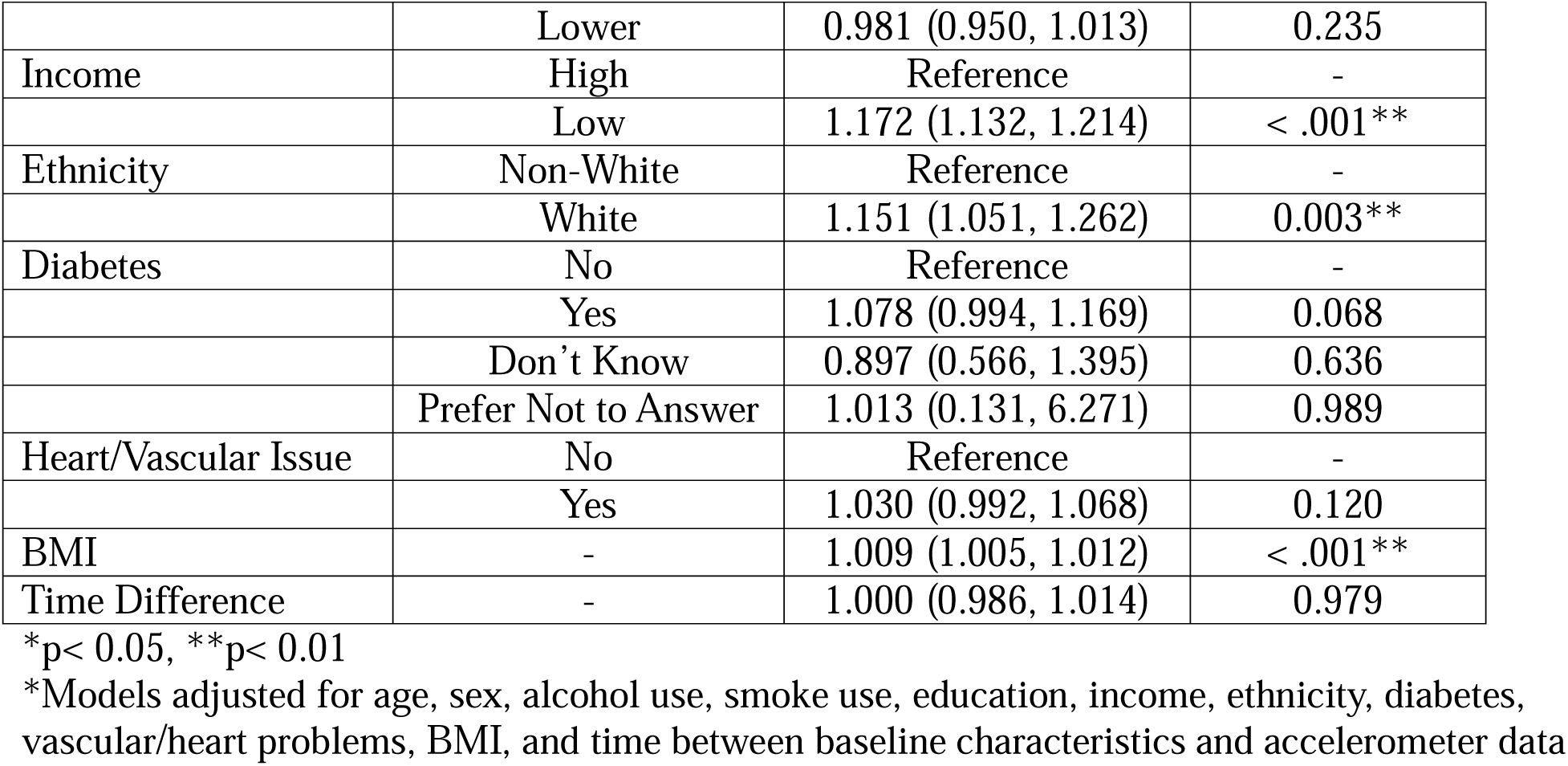
Logistic Regression Summary for Hearing/Speech-in-Noise Outcomes and % of Time Spent in Moderate-to-Vigorous Physical Activity.

### Associations of Hearing and Speech-in-Noise Outcomes with Spending at Least 150 Minutes per Week in MVPA

Spending at least 150 minutes per week in MVPA was significantly associated with lower odds of reporting a hearing problem after adjusting for the covariates in Model 3 (OR= 0.958, 95% CI [0.923, 0.995], p= 0.025) as presented in Table 2. Spending at least 150 minutes per week in MVPA reduces the odds of reporting a hearing problem by 4.20%. Similarly, spending at least 150 minutes per week in MVPA was also significantly associated with lower odds of reporting a speech-in-noise problem after adjusting for these covariates in Model 4 (OR= 0.953, 95% CI [0.922, 0.986], p= 0.005) as presented in Table 2. Spending at least 150 minutes per week in MVPA reduces the odds of reporting a hearing problem by 4.70%.

**Table 2.** Logistic Regression Summary for Hearing/Speech-in-Noise Outcome Meeting 150 Minutes of Moderate-to-Vigorous Physical Activity or Not.

| **Hearing Outcome Models** |  |  |  |
|---|---|---|---|
| Predictor | Category | OR (95% CI) | P-value |
| WHO Guidelines | Not Meet | Reference | - |
|  | Meet | 0.958 (0.923, 0.995) | 0.025* |
| Age | - | 1.049 (1.047, 1.052) | < .001** |
| Sex | Female | Reference | - |
|  | Male | 1.589 (1.535, 1.644) | < .001** |
| Alcohol Use | Never | Reference | - |
|  | Current | 1.031 (0.927, 1.149) | 0.571 |
|  | Previous | 1.143 (0.990, 1.321) | 0.069 |
|  | Prefer Not to Answer | 1.208 (0.427, 2.991) | 0.698 |
| Smoke Use | Never | Reference | - |
|  | Current | 1.192 (1.115, 1.273) | < .001** |
|  | Previous | 1.168 (1.127, 1.210) | < .001** |
|  | Prefer Not to Answer | 1.034 (0.679, 1.536) | 0.873 |
| Education | Higher | Reference | - |
|  | Lower | 0.997 (0.962, 1.032) | 0.856 |
| Income | High | Reference | - |
|  | Low | 1.109 (1.066, 1.153) | < .001** |
| Ethnicity | Non-White | Reference | - |
|  | White | 1.408 (1.261, 1.575) | < .001** |
| Diabetes | No | Reference | - |
|  | Yes | 1.057 (0.969, 1.152) | 0.210 |
|  | Don't Know | 0.981 (0.591, 1.573) | 0.938 |
|  | Prefer Not to Answer | 1.536 (0.191, 9.618) | 0.645 |
| Heart/Vascular Issue | No | Reference | - |
|  | Yes | 1.017 (0.977, 1.059) | 0.408 |
| BMI | - | 1.018 (1.012, 1.021) | < .001** |
| Time Difference | - | 0.962 (0.945, 0.977) | < .001** |
| **Speech-in-Noise Outcome Model** |  |  |  |
| Predictor | Category | OR (95% CI) | P-value |
| WHO Guidelines | Not Meet | Reference | - |
|  | Meet | 0.953 (0.922, 0.986) | 0.005** |
| Age | - | 1.036 (1.034, 1.038) | < .001** |
| Sex | Female | Reference | - |
|  | Male | 1.602 (1.542, 1.640) | < .001** |
| Alcohol Use | Never | Reference | - |
|  | Current | 1.051 (0.956, 1.157) | 0.302 |
|  | Previous | 1.280 (1.125, 1.456) | < .001** |
|  | Prefer Not to Answer | 1.592 (0.667, 3.692) | 0.281 |
| Smoke Use | Never | Reference | - |
|  | Current | 1.84 (1.115, 1.257) | < .001** |
|  | Previous | 1.177 (1.140, 1.216) | < .001** |
|  | Prefer Not to Answer | 0.907 (0.616, 1.314) | 0.612 |
| Education | Higher | Reference | - |
|  | Lower | 0.981 (0.950, 1.012) | 0.229 |
| Income | High | Reference | - |
|  | Low | 1.172 (1.132, 1.213) | < .001** |
| Ethnicity | Non-White | Reference | - |
|  | White | 1.150 (1.050, 1.261) | 0.003** |
| Diabetes | No | Reference | - |
|  | Yes | 1.077 (0.993, 1.168) | 0.071 |
|  | Don't Know | 0.896 (0.565, 1.394) | 0.633 |
|  | Prefer Not to Answer | 0.997 (0.129, 6.184) | 0.997 |
| Heart/Vascular Issue | No | Reference | - |
|  | Yes | 1.030 (0.993, 1.069) | 0.115 |
| BMI | - | 1.009 (1.005, 1.012) | < .001** |
| Time Difference | - | 1.000 (0.986, 1.014) | 0.987 |

\*p< 0.05, \*\*p< 0.01
\*Models adjusted for age, sex, alcohol use, smoke use, education, income, ethnicity, diabetes, vascular/heart problems, BMI, and time between baseline characteristics and accelerometer data

## Discussion

In a sample of 79,286 participants from UK Biobank, we investigated the association between time spent in MVPA and self-reported problems with hearing and understanding speech in noise. We found that spending more time in MVPA was associated with significantly lower odds of both reporting a hearing problem and reporting a speech-in-noise problem. We also found that meeting the World Health Organization guidelines of 150 minutes per week of MVPA was associated with significantly lower odds of both reporting a hearing problem and reporting a speech-in-noise problem. Below we discuss the implications of these findings and contextualize them within the broader research and clinical landscape.

While most of the work in this topic has been done investigating the relationship between hearing and physical activity, struggling to understand speech in noise remains the most common issue reported by those being treated for hearing loss (Billings et al., 2024; Kochkin, 2010). Yet, there has been insufficient prior work examining how physical activity is associated with speech-in-noise perception (Martinez-Amezcua et al., 2023). Given that speech-in-noise has been shown to rely on executive functions and physical activity have been proven to improve these functions, we aimed to investigate whether speech-in-noise problems were associated with physical activity.

While cognition was not directly evaluated in the current study, previous research highlighting how speech-in-noise relies on executive functions and physical activity improves these same processes strengthens our findings. Long-term exercise improves connectivity and neural efficiency in the prefrontal cortex and leads to improved executive function driven by an improvement in cardiovascular fitness (Cabral et al., 2019; Voelcker-Rehage et al., 2011). Engaging in physical activity throughout the lifespan has positive effects on maintaining cognitive health and improving brain function, with the largest effects seen in executive function (Erickson et al., 2015). Similarly, prior research also highlights the role of the frontal lobes when understanding acoustically challenging speech (Davis & Johnsrude, 2003b) as it requires increased listening effort, attention, and cognitive processes (Gosselin & Gagné, 2011; Peelle, 2018; Vaden et al., 2013; Wong et al., 2009). Specifically, performance on executive function tasks often correlates with performance on speech-in-noise tasks (Can et al., 2026; Dryden et al., 2017; Ward et al., 2016). Given that physical activity has moderate positive effects on executive functions (Singh et al., 2025) and speech-in-noise tasks rely on executive resources, we speculated that physical activity may help support speech-in-noise comprehension.

Our findings also corroborate previous research suggesting that there is a relationship between the odds of reporting a hearing problem and time spent being physically active such that those who are less physically active are more likely to report hearing problems (Gispen et al., 2014; Li et al., 2025; Martinez-Amezcua et al., 2023). Moderate hearing loss is associated with greater odds of spending less time being active (Choi et al., 2016; Gispen et al., 2014) and those who engage in MVPA compared to those who do not are at lower risk for developing hearing issues in later years (Kawakami et al., 2022; Kuo et al., 2021). Yet, it is also important to acknowledge that these are observational findings and others have found the opposite directionality of the relationship where hearing loss results in less physical activity (Martinez-Amezcua et al., 2021). Some researchers have found that those with moderate or greater hearing loss were more likely to be insufficiently active (Assi et al., 2024). While more work is needed to tease out the causal effects of the relationship, the current study expands on these findings using a larger population-based sample and objective accelerometer data illustrating that more time spent in MVPA is associated with an outcome of lower odds of reporting a hearing problem.

While the effect sizes for our models were modest, they represent an important step in identifying a meaningful relationship between hearing outcomes and accelerometer-derived MVPA in a large population-based cohort. Most novelly, they are the first to establish an association between time spent in MVPA and understanding speech in noise. This also aligns with previous work with physical activity’s effect on cognition. Physical activity has small but meaningful selective effects on cognition, with the most significant benefits on executive processes (Etnier et al., 1997). We would expect that our effect sizes would be larger given objective measures of hearing as other researchers have only seen a relationship between cognition and speech-in-noise performance for objective not self-report measures (Hoff et al., 2023). The fact that a relationship was found with subjective hearing measures is a promising and meaningful start. Our current findings open the door to future research investigating the association between speech-in-noise outcomes and physical activity with more comprehensive assessments of hearing as well as targeted interventions.

Beginning to understand how physical activity relates to speech-in-noise comprehension in the general population is an important step towards mitigating the social, cognitive, mental, and economic burdens that stem from hearing health issues. Our study was the first to directly investigate the relationship between spending time in physical activity and reporting speech-in-noise problems in a comprehensive population dataset. We found that spending more time in MVPA was significantly associated with lower odds of reporting a speech-in-noise and hearing problem which novelly advances our understanding of the association of physical activity with overall hearing health.

### Implications

The World Health Organization embraces the idea that how we hear in the future depends highly on the practices we currently take to protect our ears (Ghebreyesus, 2024; World Health Organization, 2021) However, the current focus of audiological care is on finding ways to improve audibility by amplifying sound or bypassing cochlear damage with innovative technology. Hearing aids, cochlear implants, and other auditory rehabilitation devices address structural and functional decline of auditory processes but cannot fix all the hardships that come with hearing loss. We show that a modifiable lifestyle behavior, physical activity, is associated with whether someone reports hearing issues. Not only has our study confirmed previous research on physical activity being associated with better hearing in a large population-based cohort, but it also uniquely highlights its association with speech-in-noise understanding. As there is currently a lack of widespread interventions that directly target speech-in-noise ability, our findings can lead to improving a common, yet undertreated issue.

It is important to note that despite the evidence for the importance of physical activity, 31% of adults worldwide—and some 53.1% of Americans (Elgaddal et al., 2022)—fail to meet the recommended guidelines for aerobic activity (World Health Organization, 2024). Filling this gap by promoting physical activity’s benefits may not just help the obvious such as cardiovascular health or metabolic health, but it may also simultaneously improve hearing health.

### Limitations

While the findings of this study have important implications for hearing health, there are a few limitations that must be addressed. First, this work is only based on associations and observations from a single week instance of MVPA as well as answers to just 2 simple questions. Further research should investigate longer periods of MVPA and more objective measures of both hearing outcomes. These findings would also be aided by longitudinal or interventional studies examining how increases in MVPA may be linked to hearing health. Second, there was a variable period of time between the baseline visits and accelerometer data. We controlled for this variability by adding a covariate for the time between visits, but further work should collect these variables within a closer timeframe. It is also important to acknowledge that although we found that spending more time in MVPA was associated with lower odds of reporting hearing issues, those who have better hearing may just be more active or social in general. Future studies should disentangle the directionality of this relationship.

There are other factors that may relate to both physical activity and hearing health that we did not control for in the statistical analysis, providing opportunities for future studies that can more clearly separate out the influence of various lifestyle factors and variables like cognition. Ideally, this could be done with more granular assessments of hearing health, which we anticipate would be more sensitive than the self-report measures we used here.

Another issue worth considering in future research relates to the language background of the participants. Although all participants in the current data are from the UK, individual language backgrounds (for example, English as a first or second language) may influence daily communication function, and is important to consider when interpreting the results. However, the degree to which this issue would impact self-report measures, as used here, is unclear.

A final caveat relates to reconciling accelerometer data with World Health Organization guidelines as using accelerometer data is not necessarily compatible with current physical activity guidelines. These recommendations were designed for activity on top of “normal” activity. Using accelerometry may lead people to believe they are exceeding guidelines as accelerometers detect all activities, including daily movement that was already factored in when creating the World Health Organizations guidelines (Thompson et al., 2016). Nonetheless, our study shows that spending at least 150 minutes per week in MVPA is associated with better hearing health.

## Conclusion

Overall, this study found that more time spent in MVPA is significantly associated with lower odds of reporting a hearing problem and reporting trouble with speech in noise. It also found that spending at least 150 minutes per week in MVPA was associated with lower odds of reporting both a hearing and speech-in-noise problem. Together, these findings suggest that MVPA may be beneficial to understanding speech in noise and hearing.

## Data Availability

Access to UK Biobank data is restricted to those with an approved project. Analysis code for the current project is available at https://osf.io/djn5m/.

