## Supplemental Table 1 for "The Relationship Between Moderate-to-Vigorous Physical Activity and Hearing Health in the UK Biobank"

All models passed assumption checks except for linearity of logit odds assumption. Models with hearing as the outcome variable (Models 1 and 3) had BMI as a predictor violating this assumption and models with a speech-in-noise problem as an outcome variable (Models 2 and 4) had BMI and age violating this assumption. We modeled these predictors using restricted cubic splines (df = 4) to include nonlinear relationships. The results are summarized in the tables below.

**Tables**

| **Predictor** | **Category** | **OR (95% CI)** | **P-value** |
| --- | --- | --- | --- |
| % of time in MVPA | - | 0.990 (0.983, 0.997) | 0.006** |
| Age | - | 1.049 (1.046, 1.052) | < .001** |
| Sex | Female | Reference | - |
|  | Male | 1.595 (1.539, 1.652) | < .001** |
| Alcohol Use | Never | Reference | - |
|  | Current | 1.032 (0.928, 1.150) | 0.565 |
|  | Previous | 1.143 (0.990, 1.320) | 0.069 |
|  | Prefer Not to Answer | 1.221 (0.432, 3.025) | 0.682 |
| Smoke Use | Never | Reference | - |
|  | Current | 1.189 (1.112, 1.271) | < .001** |
|  | Previous | 1.168 (1.127, 1.210) | < .001** |
|  | Prefer Not to Answer | 1.033 (0.678, 1.535) | 0.876 |
| Education | Higher | Reference | - |
|  | Lower | 0.996 (0.961, 1.031) | 0.810 |
| Income | High | Reference | - |
|  | Low | 1.109 (1.066, 1.153) | < .001** |
| Ethnicity | Non-White | Reference | - |
|  | White | 1.411 (1.264, 1.579) | < .001** |
| Diabetes | No | Reference | - |
|  | Yes | 1.057 (0.969, 1.153) | 0.207 |
|  | Don’t Know | 0.979 (0.590, 1.5730 | 0.933 |
|  | Prefer Not to Answer | 1.588 (0.204, 9.923) | 0.619 |
| Heart/Vascular Issue | No | Reference | - |
|  | Yes | 1.015 (0.975, 1.056) | 0.469 |
| BMI | rcs(BMI, 4)BMI | 0.998 (0.979, 1.018) | 0.853 |
|  | rcs(BMI, 4)BMI' | 1.103 (1.016, 1.196) | 0.019* |
|  | rcs(BMI, 4)BMI'' | 0.759 (0.607, 0.950) | 0.016* |
| Time Difference | - | 0.962 (0.947, 0.978) | < .001** |

**Supplemental Table 1.** Logistic Regression Summary for Hearing Outcome % of Time Spent in Moderate-to-Vigorous Physical Activity

*p< 0.05, **p< 0.01

*Models adjusted for age, sex, alcohol use, smoke use, education, income, ethnicity, diabetes, vascular/heart problems, BMI, and time between baseline characteristics and accelerometer data

**Supplemental Table 2.** Logistic Regression Summary for Speech-in-Noise Outcome % of Time Spent in Moderate-to-Vigorous Physical Activity

| **Predictor** | **Category** | **OR (95% CI)** | **P-value** |
| --- | --- | --- | --- |
| % of time in MVPA | - | 0.991 (0.985, 0.998) | 0.009** |
| Age | rcs(recru_age, 4)recru_age | 1.048 (1.039, 1.057) | < .001** |
|  | rcs(recru_age, 4)recru_age' | 0.967 (0.948, 0.987) | 0.001** |
|  | rcs(recru_age, 4)recru_age'' | 1.172 (1.074, 1.279) | < .001** |
| Sex | Female | Reference | - |
|  | Male | 1.603 (1.553, 1.655) | < .001** |
| Alcohol Use | Never | Reference | - |
|  | Current | 1.054 (0.958, 1.160) | 0.281 |
|  | Previous | 1.281 (1.127, 1.458) | < .001** |
|  | Prefer Not to Answer | 1.614 (0.677, 3.743) | 0.266 |
| Smoke Use | Never | Reference | - |
|  | Current | 1.186 (1.117, 1.259) | < .001** |
|  | Previous | 1.180 (1.143, 1.219) | < .001** |
|  | Prefer Not to Answer | 0.911 (0.619, 1.320) | 0.627 |
| Education | Higher | Reference | - |
|  | Lower | 0.981 (0.951, 1.013) | 0.245 |
| Income | High | Reference | - |
|  | Low | 1.174 (1.134, 1.216) | < .001** |
| Ethnicity | Non-White | Reference | - |
|  | White | 1.152 (1.052, 1.263) | 0.002** |
| Diabetes | No | Reference | - |
|  | Yes | 1.071 (0.987, 1.161) | 0.097 |
|  | Don’t Know | 0.891 (0.562, 1.386) | 0.615 |
|  | Prefer Not to Answer | 0.994 (0.128, 6.158) | 0.995 |
| Heart/Vascular Issue | No | Reference | - |
|  | Yes | 1.030 (0.992, 1.068) | 0.119 |
| BMI | rcs(BMI, 4)BMI | 0.998 (0.982, 1.015) | 0.848 |
|  | rcs(BMI, 4)BMI' | 1.024 (0.952, 1.100) | 0.526 |
|  | rcs(BMI, 4)BMI'' | 0.961 (0.788, 1.173) | 0.696 |
| Time Difference | - | 1.000 (0.986, 1.014) | 0.976 |

*p< 0.05, **p< 0.01

*Models adjusted for age, sex, alcohol use, smoke use, education, income, ethnicity, diabetes, vascular/heart problems, BMI, and time between baseline characteristics and accelerometer data

**Supplemental Table 3.** Logistic Regression Summary for Hearing Outcome Meeting 150 Minutes of Moderate-to-Vigorous Physical Activity or Not

| **Predictor** | **Category** | **OR (95% CI)** | **P-value** |
| --- | --- | --- | --- |
| WHO Guidelines | Not Meet | Reference | - |
|  | Meet | 0.959 (0.924, 0.995) | 0.027* |
| Age | - | 1.049 (1.047, 1.052) | < .001** |
| Sex | Female | Reference | - |
|  | Male | 1.587 (1.533, 1.644) | < .001** |
| Alcohol Use | Never | Reference | - |
|  | Current | 1.033 (0.929, 1.151) | 0.554 |
|  | Previous | 1.144 (0.990, 1.321) | 0.068 |
|  | Prefer Not to Answer | 1.210 (0.428, 2.995) | 0.696 |
| Smoke Use | Never | Reference | - |
|  | Current | 1.190 (1.113, 1.271) | < .001** |
|  | Previous | 1.167 (1.126, 1.209) | < .001** |
|  | Prefer Not to Answer | 1.032 (0.678, 1.534) | 0.879 |
| Education | Higher | Reference | - |
|  | Lower | 0.996 (0.962, 1.032) | 0.827 |
| Income | High | Reference | - |
|  | Low | 1.109 (1.066, 1.153) | < .001** |
| Ethnicity | Non-White | Reference | - |
|  | White | 1.409 (1.262, 1.577) | < .001** |
| Diabetes | No | Reference | - |
|  | Yes | 1.058 (0.970, 1.153) | 0.203 |
|  | Don’t Know | 0.979 (0.590, 1.570) | 0.932 |
|  | Prefer Not to Answer | 1.559 (0.200. 9.763) | 0.634 |
| Heart/Vascular Issue | No | Reference | - |
|  | Yes | 1.016 (0.976, 1.057) | 0.446 |
| BMI | rcs(BMI, 4)BMI | 0.998 (0.979, 1.018) | 0.873 |
|  | rcs(BMI, 4)BMI' | 1.103 (1.017, 1.197) | 0.018* |
|  | rcs(BMI, 4)BMI'' | 0.757 (0.605, 0.947) | 0.015* |
| Time Difference | - | 0.962 (0.947, 0.978) | < .001** |

*p< 0.05, **p< 0.01

*Models adjusted for age, sex, alcohol use, smoke use, education, income, ethnicity, diabetes, vascular/heart problems, BMI, and time between baseline characteristics and accelerometer data

**Supplemental Table 4.** Logistic Regression Summary for Speech-in-Noise Outcome Meeting 150 Minutes of Moderate-to-Vigorous Physical Activity or Not

| **Predictor** | **Category** | **OR (95% CI)** | **P-value** |
| --- | --- | --- | --- |
| WHO Guidelines | Not Meet | Reference | - |
|  | Meet | 0.955 (0.924, 0.988) | 0.008** |
| Age | rcs(recru_age, 4)recru_age | 1.048 (1.038, 1.057) | < .001** |
|  | rcs(recru_age, 4)recru_age' | 0.968 (0.948, 0.987) | 0.001** |
|  | rcs(recru_age, 4)recru_age'' | 1.171 (1.073, 1.278) | < .001** |
| Sex | Female | Reference | - |
|  | Male | 1.600 (1.550, 1.651) | < .001** |
| Alcohol Use | Never | Reference | - |
|  | Current | 1.056 (0.960, 1.162) | 0.267 |
|  | Previous | 1.282 (1.127, 1.459) | < .001** |
|  | Prefer Not to Answer | 1.601 (0.671, 3.712) | 0.275 |
| Smoke Use | Never | Reference | - |
|  | Current | 1.185 (1.116, 1.258) | < .001** |
|  | Previous | 1.180 (1.142, 1.218) | < .001** |
|  | Prefer Not to Answer | 0.910 (0.619, 1.319) | 0.625 |
| Education | Higher | Reference | - |
|  | Lower | 0.981 (0.951, 1.013) | 0.239 |
| Income | High | Reference | - |
|  | Low | 1.174 (1.133, 1.215) | < .001** |
| Ethnicity | Non-White | Reference | - |
|  | White | 1.151 (1.051, 1.262) | 0.003** |
| Diabetes | No | Reference | - |
|  | Yes | 1.070 (0.987, 1.160) | 0.100 |
|  | Don’t Know | 0.891 (0.561, 1.385) | 0.613 |
|  | Prefer Not to Answer | 0.979 (0.126, 6.078) | 0.982 |
| Heart/Vascular Issue | No | Reference | - |
|  | Yes | 1.030 (0.993, 1.069) | 0.114 |
| BMI | rcs(BMI, 4)BMI | 0.998 (0.982, 1.016) | 0.856 |
|  | rcs(BMI, 4)BMI' | 1.024 (0.953, 1.100) | 0.523 |
|  | rcs(BMI, 4)BMI'' | 0.960 (0.787, 1.171) | 0.686 |
| Time Difference | - | 1.000 (0.986, 1.014) | 0.982 |

*p< 0.05, **p< 0.01

*Models adjusted for age, sex, alcohol use, smoke use, education, income, ethnicity, diabetes, vascular/heart problems, BMI, and time between baseline characteristics and accelerometer data
